# Clinical features and laboratory inspection of novel coronavirus pneumonia (COVID-19) in Xiangyang, Hubei

**DOI:** 10.1101/2020.02.23.20026963

**Authors:** Weiliang Cao, Li Shi, Lin Chen, Xuemei Xu, Zirong Wu

**Affiliations:** Clinical Laboratory of Xiangyang No 1 Hospital, The Fourth affiliated Hospital of Hubei medicine university, xiangyang, Hubei

**Keywords:** novel coronavirus, pneumonia, COVID-19, risk factor

## Abstract

**Background:** Since December 2019, a novel coronavirus pneumonia (COVID-19) rapidly spread in China, reached multiple continents currently.We aimed to reveal the infectious characteristics of COVID-19 that provide more information for the research of novel coronavirus.

**Methods:** We performed a retrospective study on the clinical characteristics of 128 COVID-19 cases with laboratory-confirmed from Xiangyang No 1 Hospitalad during January 2020 to 16 February 2020.

**Results:** Female patients account for 53.1%. The aged below 20 years that accounts for 1.6% of overall patients. The aged in 21∼50, 51∼65, over 66 years were accounts for 44.5%, 35.1%,18.8%, respectively. In the difference age spectrum, all severe groups compared with non-severe groups were difference significantly (P < 0.01). Fever (89.8%) and Cough (67.2%) were common clinical symptoms. The rate of patients with sore throats (14.1%) was rare. The rate of chest computed tomography scan showing ground glass opacity in overall, non-severe, severe groups were 63.3%, 60.7%, 76.2%, respectively. White blood cell counts in the normal range of overall patients, but severe group patients were increased significantly (*P* < 0.01). Lymphocytes of overall patients were decreased. Alanine transaminase (ALT) and aspartate transaminase (AST) in the normal range of overall patients, but its were elevated in the severe group. Creatinine (CR) and blood urea nitrogen (BUN) of overall patients in the normal range. C-reactive protein (CRP) level of all patients were increased markedly, but it in the severe group was significantly higher than that in the non-severe group (*P* < 0.01).

**Conclusions:** Our data provide more information that advanced age, lower lymphocytes levels at the diagnosed COVID-19 patients may be a risk factor for unfavourable prognosis. The white blood cells and C-reactive protein level elevated in severe COVID-19 patients may be accompanying bacterial infection. 2019-nCov may be carries a risk factor of impaired liver and kidney function.

## INTRODUCTION

Since December 2019, fast-multiplying outbreak of the 2019 novel coronavirus (2019-nCov) in China, which have reached multiple continents currently. 2019-nCov mainly cause pneumonia that the desease was named as novel coronavirus pneumonia (COVID-19) ^1^. The 2019-nCov is an betacoronavirus that possibly originated from wild animals that genome is highly homologous to bats ^2^. Human-to-human transmission is the main way of cause infection ^3^. The World Health Organization (WHO) has declared it to be a public health emergency of international concern ^4^. Until February 20, 2020, a total number of 75569 COVID-19 cases have been diagnosed in China, including 2239 deaths ^5^. Despite the rapid spread over the world, the clinical features and laboratory inspection of COVID-19 keep largely unclear. The clinical characteristics of severe COVID-19 cases were similar to severe acute respiratory syndrome (SARS) and the Middle East Respiratory Syndrome (MERS) that could occur acute respiratory distress syndrome (ARDS), acute cardiac injury, and even death ^6^.Whereas the main clinical features of COVID-19 patients are fever, cough and sore throat. However, there are few reports on the clinical features and laboratory detection results of NCP cases in every regions of Hubei province outside Wuhan. We aimed to through analyzed 128 laboratory confirmed cases of Xiangyang No. 1 Hospital to reveal the infectious characteristics of COVID-19 that provide more information for the research of novel coronavirus.

## METHODS

We performed a retrospective study on the clinical characteristics of laboratory-confirmed 128 cases with COVID-19. Patients were admitted to the Xiangyang No.1 Hospital from 1 January 2020 to 16 February 2020, with final follow-up for the research on 21 February 2020. Clinical data were collected from the hospital electronic medical records system and laboratory information system. On admission, 107 and 21 patients were divided into non-severe and severe groups, respectively. A confirmed case with COVID-19 was defined as a positive result to real-time reverse transcriptase polymerase chain reaction (RT-PCR) assay for nasal and pharyngeal swab specimens ^7^. Only the laboratory-confirmed cases were included the analysis. Suspected patients and clinical diagnosised patients were not part of our study. The study was approved by the ethics committee of the Xiangyang No. 1Hospital with written informed consent from the patients.

All statistical analyses were performed using SPSS version 20.0 (IBM, Armonk, NY, USA). The data between groups with variables were compared by χ2 test or Fisher’s exact test, and one-way analysis of variance followed by Tukey’s test. All means were reported with the corresponding standard deviation in tables. The *P* values < 0.05 were regarded as statistically significant.

## RESULTS

### Clinical features and Demographic

The 128 COVID-19 patients were confirmed by laboratory detection. On admission, 107 and 21 patients were divided into non-severe and severe groups, respectively. The demographic and clinical features were showed in **Table 1**. The data showed that female patients account for 53.1%, the proportion of these female patients in the severe and non-severe groups was 55.1% and 42.9%, respectively. The overall patients throughout the whole spectrum of age. The aged below 20 years that accounts for 1.6% of overall patients. The rate of overall patients who aged in 21∼50 years were 44.5%, no-severe and severe patients accounts for 46.7%, 33.3%, respectively. The 51∼65 years accounts for 35.1% of overall patients. The older patients aged over 66 years in the group of overall patients accounts for 18.8%. However, aged over 50 years accounting for 53.9% of overall patients. In the difference age spectrum, all severe groups compared with non-severe groups were difference significantly (*P* < 0.01). Fever (89.8%) and Cough (67.2%) were common clinical symptoms. The rate of patients with sore throats (14.1%) was rare. The rate of chest computed tomography scan showing ground glass opacity in overall, non-severe, severe were 63.3%, 60.7%, 76.2%, respectively. However, no marked differences in fever, cough, sore throat between the two groups (all *P* > 0.05). Moreover, there were distinctly different of chest computed tomography scan showing ground glass opacity in severe group compared with non-severe group (*P* < 0.05).

**Table 1.**
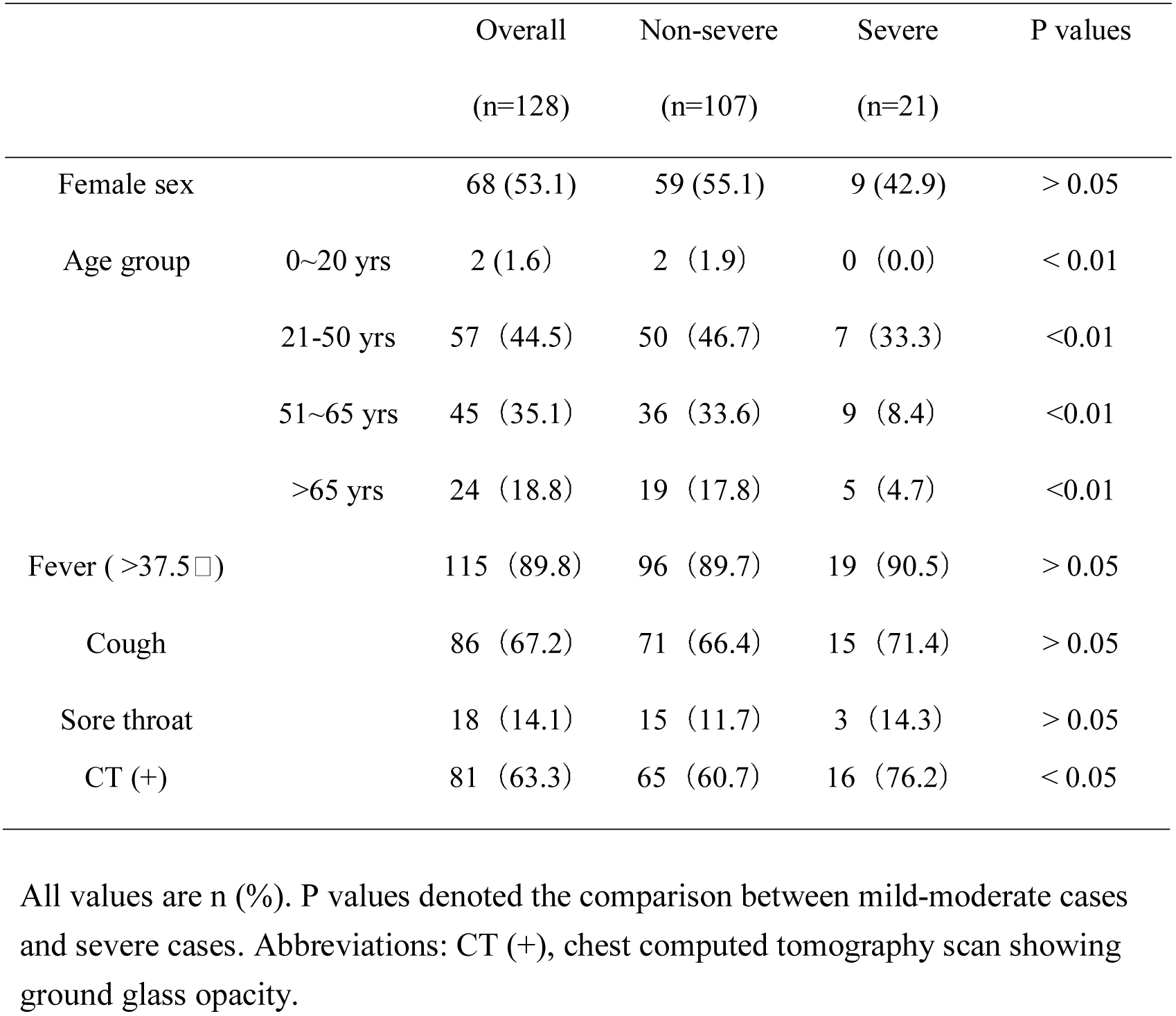
Clinical features and Demographic of COVID-19 patients (n=128).

### Laboratory inspection of patients infected 2019-nCov

All patients were summoned to the laboratory for a routine blood sample, all of which were analyzed. Laboratory detection results were showed in **Table 2**. Results indicated that white blood cell counts in the normal range of overall patients, but severe group patients were increased significantly (*P* < 0.01). Lymphocytes of overall patients were decreased, yet Platelets in the normal range. Whereas, Lymphocytes and Platelets level of severe group obviously lower than no-severe group (*P* < 0.01). Alanine transaminase and aspartate transaminase in the normal range of overall patients, but its were elevated in the severe group. Creatinine and blood urea nitrogen of overall patients in the normal range and no marked difference between no-severe and severe group. C-reactive protein level of all patients were increased markedly, but it in the severe group was significantly higher than that in the non-severe group (*P* <0.01).

**Table 2.**
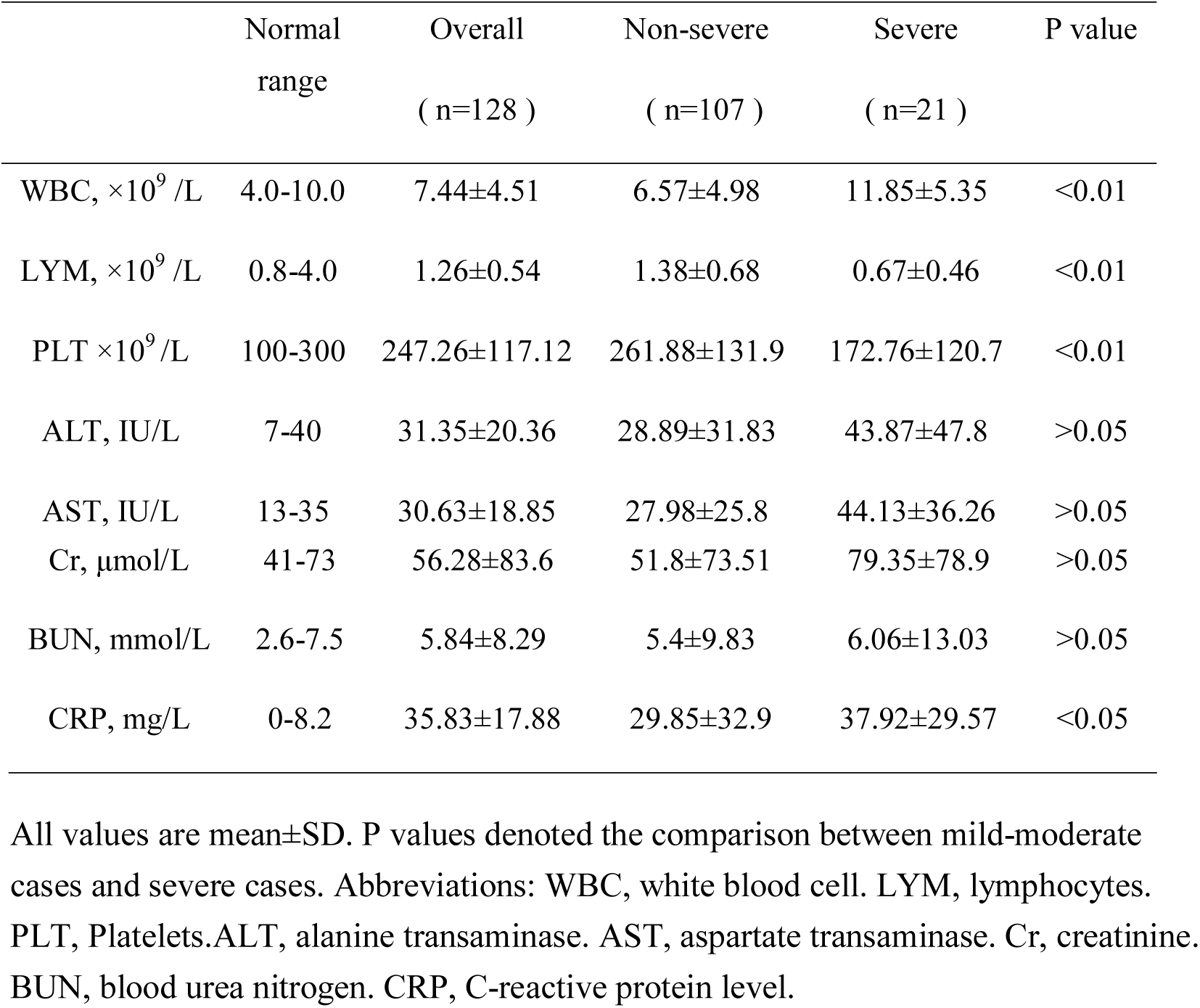
Laboratory inspection of COVID-19 patients (n=128).

## DISCUSSION

2019 novel coronavirus was an batacoronavirus which was transmitted by droplets and direct contact with a patient’s body fluids, other ways of it transmission was unclear^8,9^. Until 16 February 2020, the number of COVID-19 patients was more than 70 000, yet the number of infections was still rising ^5^. However, there are still no effective treatment for COVID-19, the main treatment is symptomatic and supportive therapy to alleviate symptoms and maintain organ function in severe diseases^10^. Although some studies have reported that some drugs can be used against the 2019-nCov, these drugs still in the research stage^11^.

Our 128 case series by laboratory-diagnosed that all of them comeback from Wuhan, or had contacted with people from Wuhan, or close contacted with diagnosed patients. However, there are unavoidable misdiagnosis through laboratory-confirmed by real-time quantitative fluorescence PCR detection of 2019-nCov due to sampling deviation. In our study, the aged over 50 years patients accounted for over half of the diagnosed patients (53.9%). Fever (89.8%) and cough(67.2%) were common symptoms, yet sore throat (14.1%) was rare. Chest computed tomography scan showing ground glass opacity of diagnosed patients account for 63.3%, whereas the severe pneumonia up to 76.2%, these were inconsistent with other studies^12^. Our data suggested that gender may not be an influence factor in 2019-nCov infection, yet advanced age was a risk factor for viral infection and poor prognosis.

Our data of laboratory detection indicated that the white blood cells and C-reactive protein level were elevated in severe case series and the lymphocytes were decreased. The results suggested that severe pneumonia may be accompanying bacterial infection and immune deficiency may be also a risk factor for unfavourable prognosis in patients^13^. In addition, alanine transaminase (ALT), aspartate transaminase (AST) and creatinine (CR) were elevated in the patients of severe pneumonia. The results suggested that 2019-nCov may be carries a risk factor of impaired liver and kidney function.

In summary, our data provide more information that advanced age, lower lymphocytes levels at the diagnosed COVID-19 patients may be a risk factor for unfavourable prognosis. The white blood cells and C-reactive protein level elevated in severe COVID-19 patients may be accompanying bacterial infection. 2019-nCov may be carries a risk factor of impaired liver and kidney function.

## Data Availability

All authors declare no competing interests of this study.

## ACKNOWLEDGEMENTS

We thank Xuemei Xu, Hao Li, Li Shi, Le Xiong, Lin Chen, Zirong Wu for supporting this work.

## DECLARATIONS

### Ethical approval

*The study was approved by the ethics committee of the Xiangyang No. 1Hospital with written informed consent from the patients*.

